# Analyzing inherent biases in SARS-CoV-2 PCR and serological epidemiologic metrics

**DOI:** 10.1101/2020.08.30.20184705

**Authors:** Monia Makhoul, Farah Abou-Hijleh, Shaheen Seedat, Ghina R Mumtaz, Hiam Chemaitelly, Houssein Ayoub, Laith J. Abu-Raddad

**Author notes:** Address reprints requests or correspondence to Professor Laith J. Abu-Raddad, Infectious Disease Epidemiology Group, World Health Organization Collaborating Centre for Disease Epidemiology Analytics on HIV/AIDS, Sexually Transmitted Infections, and Viral Hepatitis, Weill Cornell Medicine - Qatar, Qatar Foundation - Education City, P.O. Box 24144, Doha, Qatar.

## Abstract

**Background:** Prospective observational data show that infected persons with the severe acute respiratory syndrome coronavirus 2 (SARS-CoV-2) remain polymerase chain reaction (PCR) positive for a prolonged duration, and that detectable antibodies develop slowly with time. We aimed to analyze how these effects can bias key epidemiological metrics used to track and monitor SARS-CoV-2 epidemics.

**Methods:** An age-structured mathematical model was constructed to simulate progression of SARS-CoV-2 epidemics in populations. PCR testing to diagnose infection and cross-sectional surveys to measure seroprevalence were also simulated. Analyses were conducted on simulated outcomes assuming a natural epidemic time course and an epidemic in presence of interventions.

**Results:** The prolonged PCR positivity biased the epidemiological measures. There was a lag of 10 days between the *true* epidemic peak and the *actually-observed* peak. Prior to epidemic peak, PCR positivity rate was 2-fold higher than that based only on current active infection, and half of those tested positive by PCR were in the prolonged PCR positivity stage after infection clearance. Post epidemic peak, PCR positivity rate poorly predicted true trend in active infection. Meanwhile, the prolonged PCR positivity did not appreciably bias estimation of the basic reproduction number *R*_0_. The time delay in development of detectable antibodies biased measured seroprevalence. The *actually-observed* seroprevalence substantially underestimated *true* prevalence of ever infection, with the underestimation being most pronounced around epidemic peak.

**Conclusions:** Caution is warranted in interpreting PCR and serological testing data, and any drawn inferences need to factor the effects of the investigated biases for an accurate assessment of epidemic dynamics.

## Background

The severe acute respiratory syndrome coronavirus 2 (SARS-CoV-2) emerged in late 2019 [1] and resulted in a pandemic [2]. As the number of laboratory-confirmed cases and coronavirus disease 2019 (COVID-19) related deaths continue to rise [2], this virus will persist as a global public health concern.

At present, the main diagnostic modality for SARS-CoV-2 is the polymerase chain reaction (PCR) test, which is typically performed on either an upper or lower respiratory tract sample. Current understanding of the infection course suggests a latent phase of few days, followed by an infectious phase also of few days before recovery [1, 3-6]. Individuals infected with the virus test positive by PCR if tested during these first two stages, but also can test positive during the recovery stage for 2-4 weeks, reflecting genetic remnants of the virus [7, 8]. The latter duration defines the *prolonged PCR positivity* duration following end of infectiousness.

Individuals recovered from this infection also typically do not develop *detectable* IgG antibodies immediately, but 2-4 weeks thereafter [7, 8]. The latter duration defines the *pre-antibody positivity* duration following end of infectiousness.

The presence of the prolonged PCR positivity duration and the pre-antibody positivity duration can complicate the epidemiological inferences drawn from population-based testing by PCR and serological assays. Using mathematical modeling simulations, the aim of the present study is to analyze how these durations can bias the key epidemiological metrics that are used to track and monitor SARS-CoV-2 epidemics, for the purpose of improving interpretation of PCR and serological testing data and our understanding of local epidemics, but also for better management of the adverse implications of the social and physical distancing restrictions.

## Methods

### Mathematical model

An age-structured mathematical model was developed to simulate SARS-CoV-2 transmission dynamics in a given generic population (Appendix Figure S1), as informed by recent modeling studies [9-11]. The model was structured factoring current understanding of SARS-CoV-2 epidemiology, and stratifies the population into compartments according to age group, infection status, infection stage, and disease stage. Following a latency period, infected individuals progress to either asymptomatic/mild infection followed by recovery, or they progress to severe or critical infection. Severe or critical infection progresses to severe or critical disease, respectively, prior to recovery, but critical disease cases have an additional risk for COVID-19 mortality.

The model further includes three tracking population compartments for the prolonged PCR positivity, pre-antibody positivity, and antibody positivity. Informed by empirical evidence [7, 8], it was assumed that infected individuals remain in the prolonged PCR positivity stage for 3 weeks on average and in the pre-antibody positivity stage also for 3 weeks on average. Some of the analyses below explored the impact of other values for these durations.

Description of the model structure, equations, and parameters are in the Appendix. All analyses were conducted on the MATLAB R2019a platform.

### Analysis scenarios

Two types of SARS-CoV-2 epidemics were simulated in this generic population: one assuming a basic reproduction number *(R*_0_) of 3.0, reflecting the natural course of the epidemic in *absence* of any social or physical distancing interventions [12, 13], and one assuming an *R*_0_ of 1.6, reflecting an epidemic in *presence* of these interventions, such as that of Qatar where *R*_0_ was estimated at about 1.6 [10].

Random PCR testing was simulated on this population through Monte Carlo sampling. Trend in PCR positive diagnoses was generated assuming first that individuals are PCR positive *only* during infection latency and infectiousness (that is during *only* active infection), and then assuming that individuals are PCR positive during infection latency, infectiousness, *and* the prolonged PCR positivity following end of infectiousness. These two simulated trends represent thus the *true* active infection presence in the population and the *actually-observed* presence through PCR testing, respectively.

Repeated *daily* cross-sectional surveys to measure antibody prevalence (seroprevalence) were also simulated on this population by Monte Carlo sampling a random sample every day. The trend in seroprevalence was generated assuming that individuals develop detectable antibodies *immediately* following onset of infection (that is detectable antibodies reflect actual infection *once* the infection occurs), and then assuming that individuals develop detectable antibodies *only* after passing through the stage of pre-antibody positivity following end of infectiousness. Once antibodies develop, it was assumed that they would persist for a long duration, beyond the simulation timeframe. These two simulated trends represent thus the *true* prevalence of ever infection in the population and the *actually-observed* seroprevalence as measured using serological assays, respectively.

## Results

Figure 1 shows the simulated daily number of PCR-positive diagnosed cases in the scenario that PCR positivity measures *true* active infection presence in the population compared to the *actually-observed* scenario in presence of the prolonged PCR positivity. There is a lag of 10 days between the *true* peak in infection incidence and the *actually-observed* peak in infection incidence when *R*_0_ is 1.6, and a lag of 5 days when *R*_0_ is 3.0. Moreover, the scenario incorporating the prolonged PCR positivity results in more cases being diagnosed than the scenario in which infected individuals are PCR positive only during active infection.

**Figure 1.**
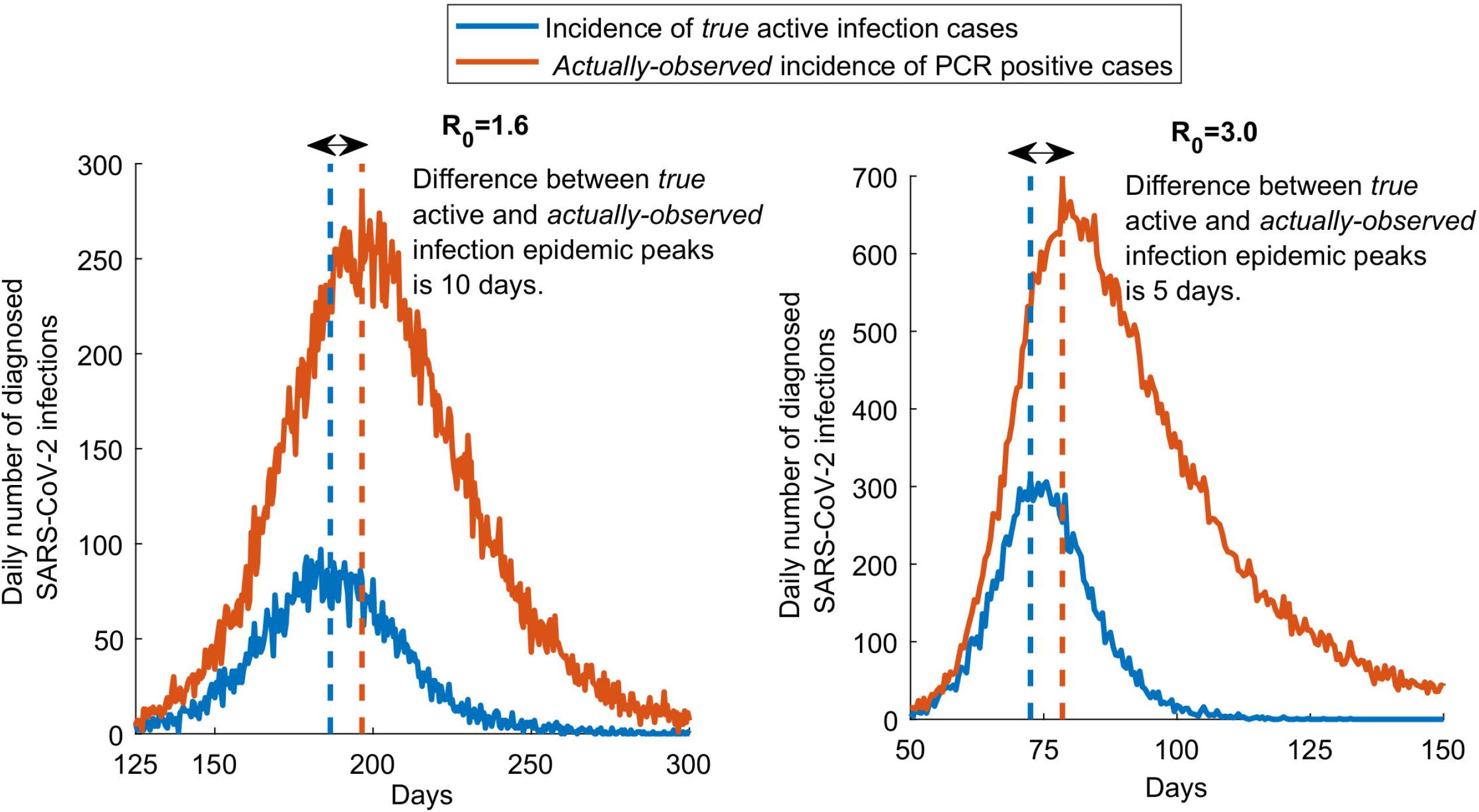
Effect of the prolonged PCR positivity on the observed trend of diagnosed cases. Daily number of new diagnosed cases of *true* active infection versus the *actually-observed* diagnosed cases in presence of the prolonged PCR positivity. The prolonged PCR positivity is assumed to last on average for three weeks after end of infectiousness [7, 8]. Two scenarios are presented, one for an *R*_0_ of 1.6 (an epidemic in presence of social and physical distancing interventions) and an *R*_0_ of 3.0 (natural course of the epidemic in absence of any social or physical distancing interventions).

Figure 2 shows the ratio of the proportion of tests that are PCR positive (“positivity rate”) in presence of the prolonged PCR positivity divided by the proportion of tests that are PCR positive assuming no prolonged PCR positivity. This ratio is shown assuming a prolonged PCR positivity duration of 2, 3, 4, or 6 weeks. Prior to the epidemic peak, the proportion of tests that are PCR positive in presence of the prolonged PCR positivity is 2-fold higher than that assuming no prolonged PCR positivity. Meanwhile, after the epidemic peak, the ratio of the two proportions steadily increases and is higher the longer is the prolonged PCR positivity—that is more and more of the infections are diagnosed not during active infection, but during the prolonged PCR positivity stage. These results were generated assuming an *R*_0_ of 1.6, and the results assuming an *R*_0_ of 3.0 show the same pattern (Appendix Figure S2).

**Figure 2.**
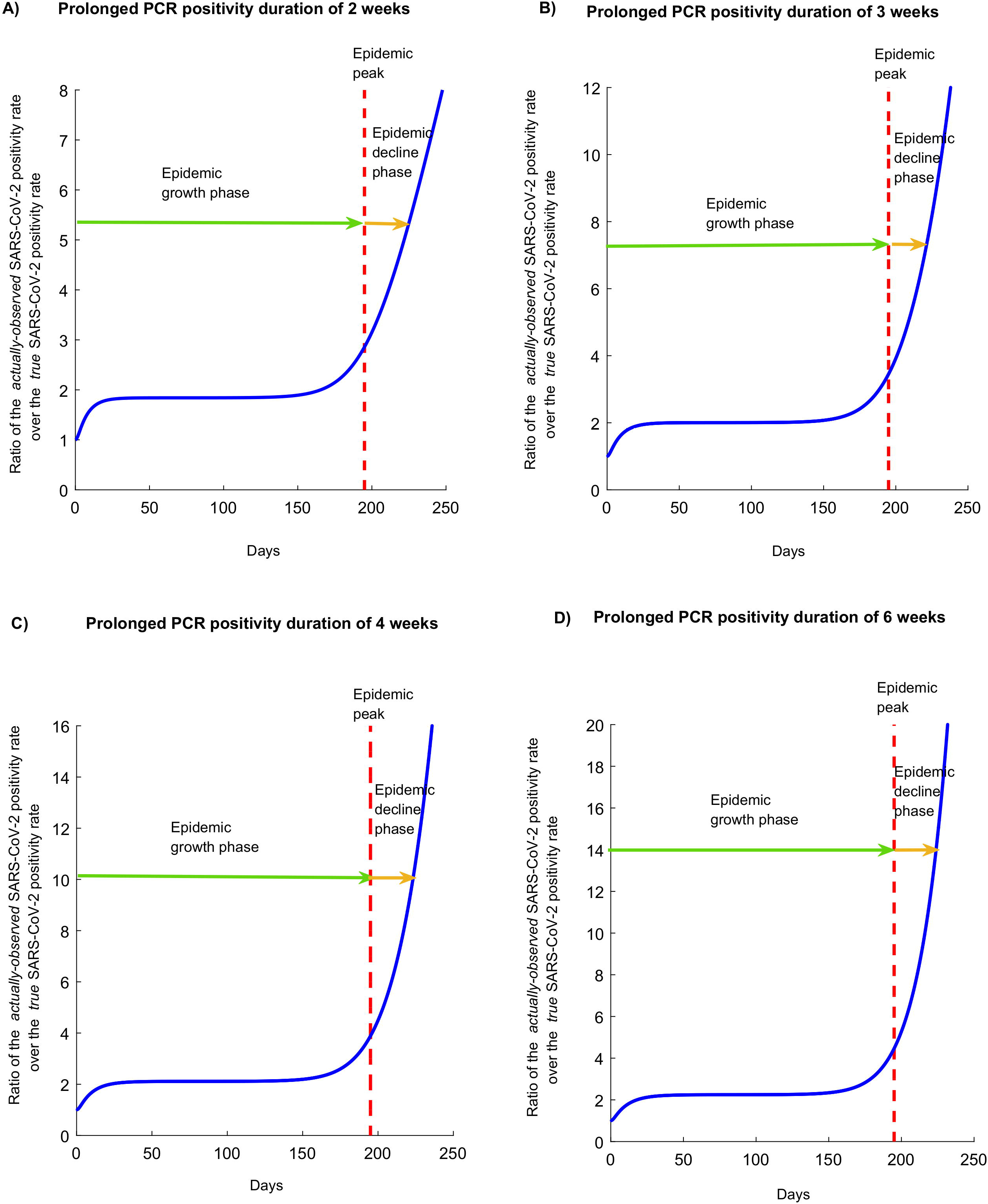
Effect of the prolonged PCR positivity on the observed SARS-CoV-2 positivity rate. Ratio of the proportion of tests that are PCR positive (“positivity rate”) in presence of the prolonged PCR positivity over the proportion of tests that are PCR positive assuming no prolonged PCR positivity. The prolonged PCR positivity is assumed to last on average for 2, 3, 4, and 6 weeks. In this epidemic simulation, *R*_0_ has a value of 1.6, that is an epidemic time course in presence of social and physical distancing interventions. The simulation for *R*_0_ of 3.0, that is for the natural course of the epidemic in absence of any social or physical distancing interventions, is found in Figure S2.

Figure 3 presents the difference in days between the epidemic peak as measured in presence of the prolonged PCR positivity and the epidemic peak based on true incidence of active infection in the population, assuming a prolonged PCR positivity duration of 2, 3, 4, or 6 weeks. The delay between the *true* epidemic peak and the *observed* epidemic peak increased as the duration of prolonged PCR positivity increased. This delay ranged from 7.5 days up to 16.5 days at an *R*_0_ of 1.6, and from 4.5 days up to 8.0 days at an *R*_0_ of 3.0.

**Figure 3.**
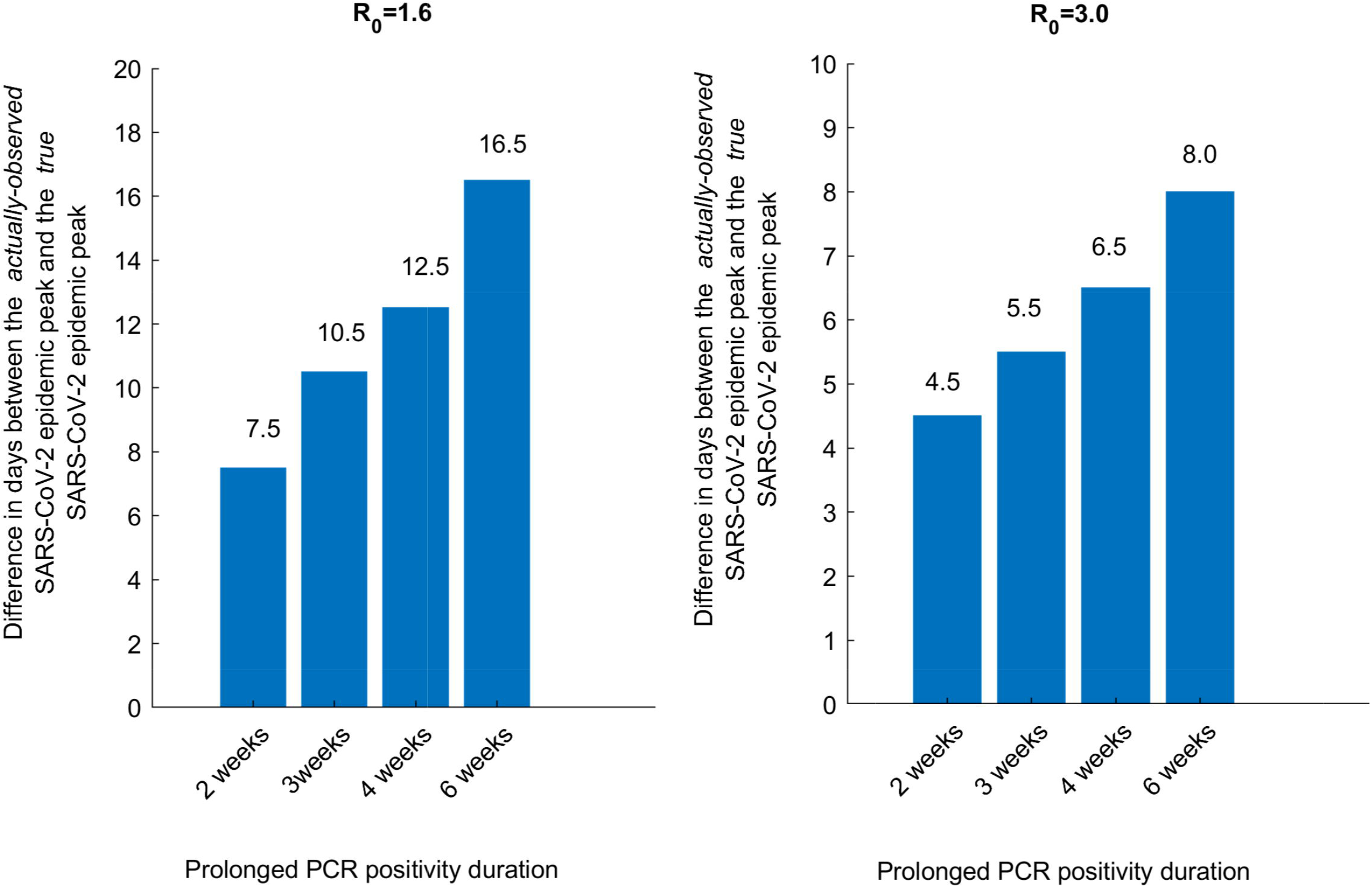
Effect of the prolonged PCR positivity on the observed SARS-CoV-2 epidemic peak. Time difference between the *actually-observed* epidemic peak in presence of the prolonged PCR positivity and the *true* epidemic peak based on true incidence of active infection in the population. The prolonged PCR positivity is assumed to last on average for 2, 3, 4, and 6 weeks. Two scenarios are presented, one for an *R*_0_ of 1.6 (an epidemic in presence of social and physical distancing interventions) and an *R*_0_ of 3.0 (natural course of the epidemic in absence of any social or physical distancing interventions).

Figure 4 and Figure S3 of the Appendix illustrate the change throughout the epidemic in the proportion of those who test positive by PCR and are latently infected, infectious, or post-infectious (that is in the prolonged PCR positivity stage) for *R*_0_ = 1.6 and *R*_0_ = 3.0, respectively.

**Figure 4.**
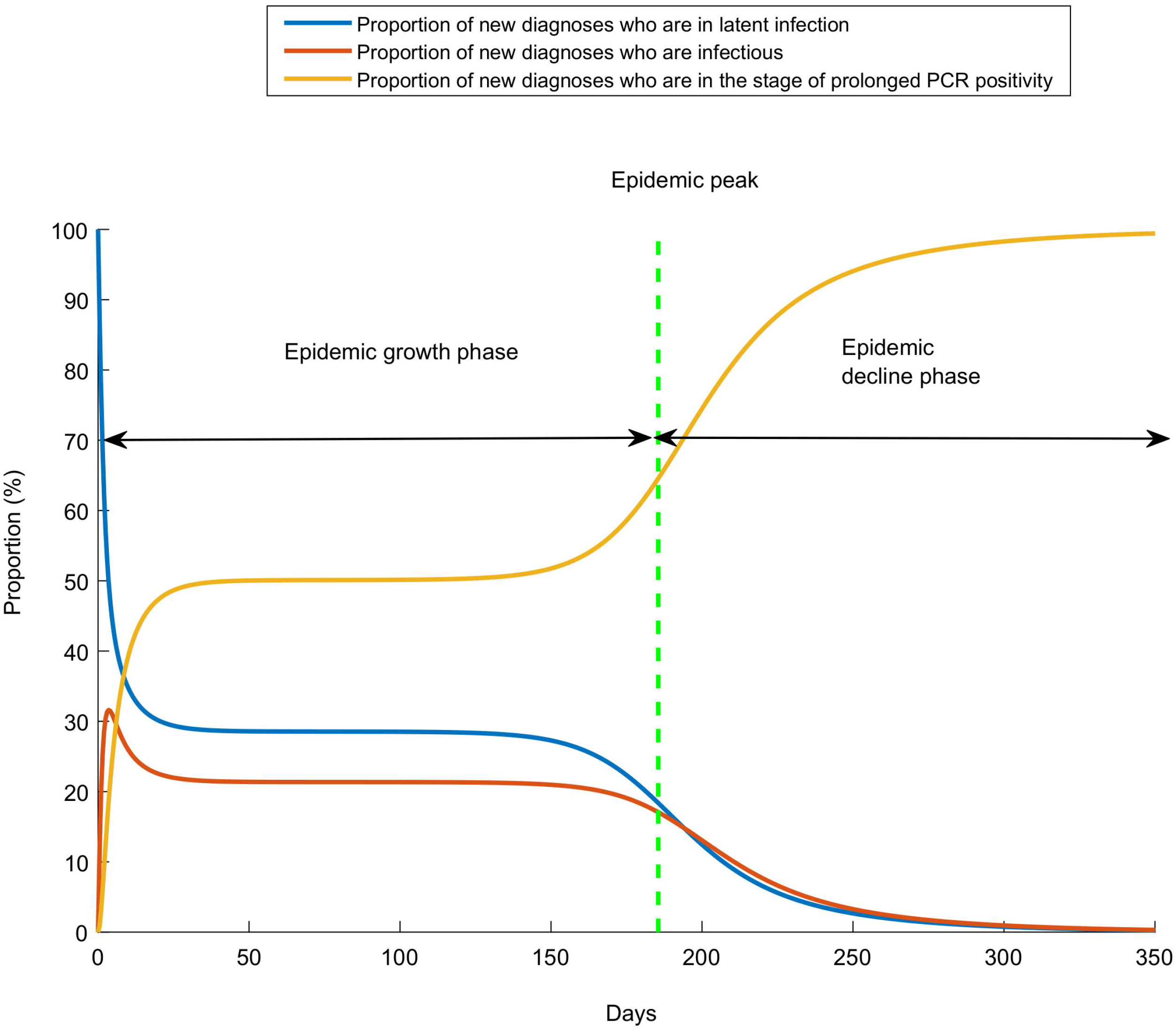
Effect of the prolonged PCR positivity on distribution of those latently infected, infectious, and post-infectious. Proportion of new diagnoses who are in latent infection, stage of infectiousness, or stage of prolonged PCR positivity. The prolonged PCR positivity is assumed to last on average for three weeks after end of infectiousness [7, 8]. In this epidemic simulation, *R*_0_ has a value of 1.6, that is an epidemic time course in presence of social and physical distancing interventions. The simulation for *R*_0_ of 3.0, that is for the natural course of the epidemic in absence of any social or physical distancing interventions, is found in Figure S3.

For *R*_0_ = 1.6, prior to the epidemic peak, approximately half of those who test positive by PCR are in the prolonged PCR positivity stage (that is already recovered from the infection). After the epidemic peak, this proportion rises steeply as the epidemic begins to decline. A similar pattern is seen for *R*_0_ *=* 3.0 (Figure S3 of the Appendix).

Figure S4 of the Appendix shows the estimated *R*_0_ as derived from the epidemic curve of diagnosed cases *in presence* and *in absence* of the prolonged PCR positivity. Two scenarios are presented, the first for an *R*_0_ of 1.6, and the second for an *R*_0_ of 3.0, and each factoring a prolonged PCR positivity duration of 2, 3, 4, or 6 weeks. The estimated *R*_0_ from the *actually-observed* diagnosed cases is always lower than that estimated from the *true* (active infection) diagnosed cases, but the difference is small, particularly so for the case of *R*_0_ of 1.6, and is not much affected by the duration of the prolonged PCR positivity.

Figure 5 shows the trend in the *true* prevalence of ever infection in the population versus the *actually-observed* seroprevalence factoring the 3 weeks average delay in the development of detectable antibodies [7, 8]. Two scenarios are presented, the first for an *R*_0_ of 1.6 and the second for an *R*_0_ of 3.0. There is a time delay in the *actually-observed* seroprevalence reaching the *true* prevalence of ever infection in the population, and this delay varies with time reflecting the epidemic phase (particularly closeness to the epidemic peak) and the intensity of the epidemic (value of *R*_0_).

**Figure 5.**
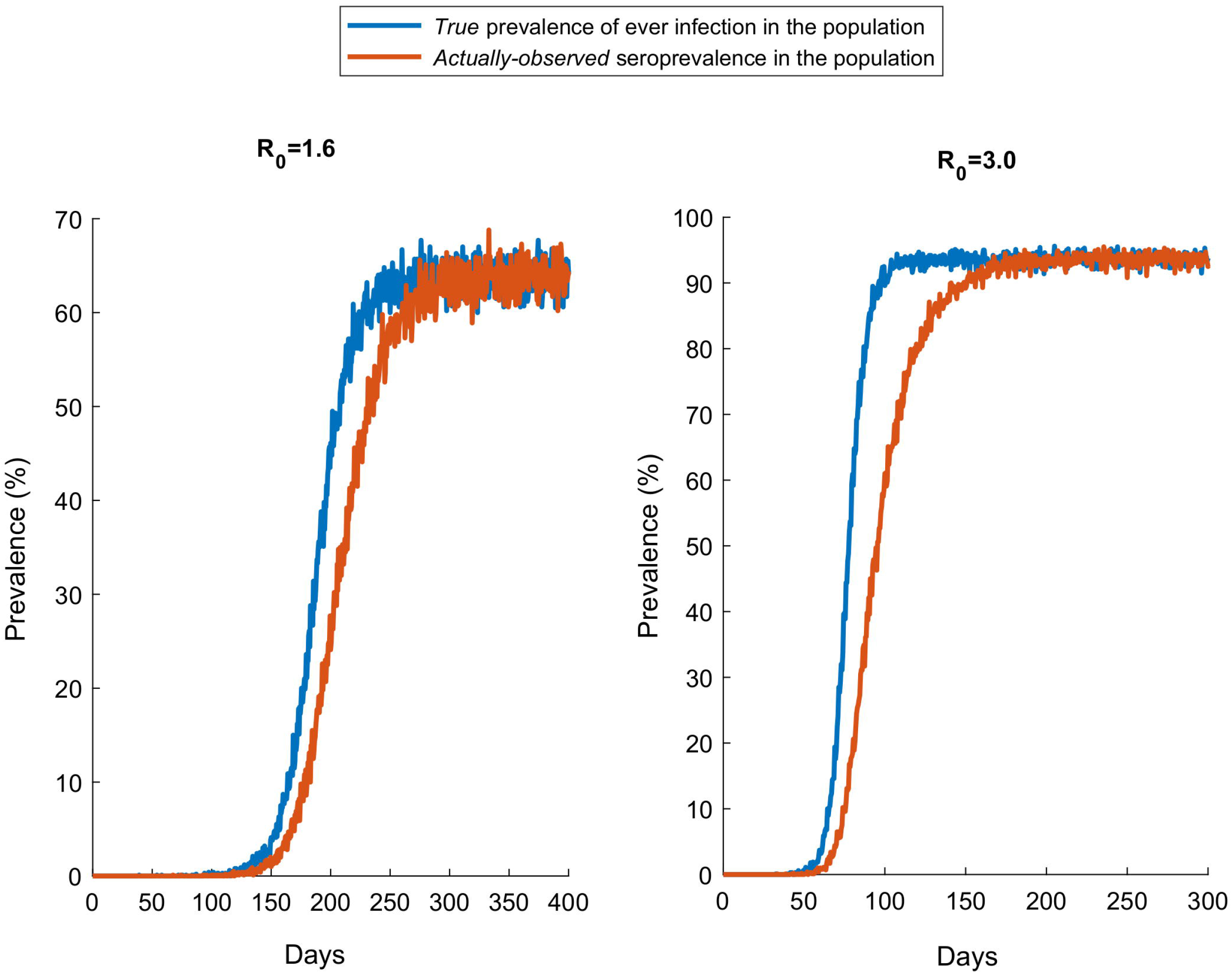
Effect of delay in development of detectable antibodies on observed seroprevalence in cross-sectional surveys. Trend in the *true* prevalence of ever infection in the population versus the *actually-observed* seroprevalence factoring the 3 weeks average delay in the development of detectable antibodies [7, 8]. Two scenarios are presented, one for an *R*_0_ of 1.6 (an epidemic in presence of social and physical distancing interventions) and an *R*_0_ of 3.0 (natural course of the epidemic in absence of any social or physical distancing interventions).

## Discussion

Management of an epidemic depends on availability of high quality real-time data in order to make the best decisions. Above results show that presence of the prolonged PCR positivity, one of the distinctive features of the SARS-CoV-2 infection reflecting the presence of genetic remnants of the virus in those who cleared their infection [7, 8], biases the epidemiological metrics and inferences drawn from the trend of PCR-positive diagnosed cases. While the prolonged PCR positivity allows more infections to be diagnosed (Figure 1), it biases assessment of the epidemic phase. The *true* phase of the epidemic (epidemic peak and also epidemic growth or decline) occurs 1-2 weeks *before* the *actually-observed* phase of the epidemic (Figures 1 and 3). This implies that the trend in PCR-positive diagnosed cases does not reflect the current status of the epidemic, but the status 1-2 weeks earlier. However, the prolonged PCR positivity does not appreciably bias the derivation of *R*_0_ from the actually-observed trend in diagnosed cases (Appendix Figure S4).

The prolonged PCR positivity also biases the testing positivity rate. The *actually-observed* positivity rate is 2-fold higher than the *true* positivity rate of active infection during the epidemic growth phase, and several folds higher during the epidemic decline phase (Figure 2). As the epidemic declines, the value of the positivity rate in conveying the actual epidemic dynamics erodes steadily with time. Moreover, during the epidemic growth phase, as much as half of those who test positive by PCR are in the prolonged PCR positivity stage having already recovered from the infection. During the epidemic decline phase, increasingly most of those testing positive are found in the prolonged PCR positivity stage and not in active infection (Figure 4). Remarkably, at all times, those newly diagnosed with the infection are likely to be found in a non-infectious stage.

Above results demonstrate that the documented time delay in development of detectable antibodies [7, 8] biases measures of seroprevalence that are derived from cross-sectional surveys of the population. At all times prior to end of the epidemic cycle, *actually-observed* seroprevalence underestimates *true* prevalence of ever infection in the population (Figure 5). The difference between what is actually observed and what is true is most pronounced around the epidemic peak. This finding demonstrates that current seroprevalence studies can significantly underestimate actual infection exposure in the population at large; an important consideration given that communities are increasingly undertaking sero-surveys to understand better virus spread and to develop efficient plans for managing the hefty costs of the social and physical distancing restrictions on society and economy.

This study has limitations. PCR testing was assumed random, but in reality this depends on the actual testing policy that is enforced in any setting. For instance, administering PCR testing to only those presenting with clinical symptoms will differentially bias detection towards those who acquired the infection within the last 5-10 days. The above results thus need to be complemented with further analysis for each specific setting to factor the actually-enforced testing policy. While the prolonged PCR positivity and the time delay in development of detectable antibodies are well-established in the literature [7, 8], it is still not sufficiently known how these are distributed in the population by age and sex, factors that may influence these effects and their impact on epidemiological measures. Epidemiological metrics can also be biased by other aspects of PCR and antibody testing, such as assay sensitivity and specificity, which are not investigated in the present study.

## Conclusions

The prolonged PCR positivity observed in SARS-CoV-2 infected persons and the time delay in development of detectable antibodies can bias key epidemiological metrics used to track and monitor SARS-CoV-2 epidemics leading to implications for managing the social and physical distancing restrictions. Caution is warranted in interpreting PCR and serological testing data, and any drawn inferences need to factor these biases for an accurate assessment of epidemic dynamics. These findings also suggest that analysis of PCR and serological testing data should not only be based on dichotomous outcomes (positive versus negative), but should also factor the quantitative measures of PCR and serological assays (such as PCR cycle threshold and antibody optical density values) to improve interpretation of these metrics.

**List of abbreviations**
SARS-CoV-2: Severe acute respiratory syndrome coronavirus 2
COVID-19: Coronavirus disease 2019
PCR: Polymerase chain reaction

## Declarations

### Ethics approval and consent to participate

This is a mathematical modelling study and all data used are aggregated, de-identified, and anonymized. Hence, this study does not need ethical consideration, informed consent of the patient or an approved protocol for the research project.

### Availability of data and materials

The model’s MATLAB codes can be obtained by contacting the authors.

## Data Availability

The MATLAB codes of the model can be obtained by contacting the authors.

## Acknowledgments

The authors are grateful for support provided by the Biomedical Research Program and the Biostatistics, Epidemiology, and Biomathematics Research Core, both at Weill Cornell Medicine-Qatar. GM acknowledges support by UK Research and Innovation as part of the Global Challenges Research Fund, grant number ES/P010873/1. The findings presented are solely the responsibility of the authors.

## Author’s contributors

MM constructed and coded the mathematical model and conducted the analyses. HA parametrized the model. FMA and LJA wrote the first draft of the paper. HC and SS supported model development and parametrization. LJA conceived and led the design of the study, construct and parameterization of the mathematical model, and drafting of the article. All authors contributed to results interpretation, discussion, and critical revision of the manuscript. All authors have read and approved the final manuscript.

## Disclose funding received for this work

Others

